# Working from home due to the COVID-19 pandemic abolished the sleep disturbance vulnerability of late chronotypes relieving their predisposition to depression

**DOI:** 10.1101/2022.02.02.22270278

**Authors:** Federico Salfi, Aurora D’Atri, Giulia Amicucci, Lorenzo Viselli, Maurizio Gorgoni, Serena Scarpelli, Valentina Alfonsi, Michele Ferrara

## Abstract

**Background and aims:** Eveningness is distinctively associated with sleep disturbances and depression symptoms due to the misalignment between biological and social clock. The widespread imposition of remote working due to the COVID-19 pandemic allowed a more flexible sleep schedule. This scenario could promote sleep and mental health of evening-type subjects. We investigated the effect of working from home on sleep quality/quantity and insomnia symptoms within the morningness-eveningness continuum, and its indirect repercussions on depressive symptomatology.

**Methods:** 610 Italian office workers (mean age ± standard deviation, 35.47 ± 10.17 yrs) and 265 remote workers (40.31 ± 10.69 yrs) participated in a web-based survey during the second contagion wave of COVID-19 (28 November–11 December 2020). We evaluated chronotype, sleep quality/duration, insomnia, and depression symptoms through validated questionnaires. Three moderated mediation models were performed, testing the mediation effect of sleep variables on the association between morningness-eveningness continuum and depression symptoms, with working modality (office vs. remote working) as moderator of the relationship between chronotype and sleep variables.

**Results:** Remote working led to delayed bedtime and get-up time. Working modality moderated the chronotype effect on sleep variables, as eveningness was related to worse sleep disturbances and shorter sleep duration only among the office workers. Working modality moderated the mediation of sleep variables between chronotype and depression. The above mediation vanished among remote workers.

**Conclusions:** Remote working strikingly abolished the vulnerability to sleep problems of evening-type subjects, relieving their predisposition to depressive symptomatology. A working environment complying with individual circadian preferences might ensure an adequate sleep quantity/quality to late chronotypes, promoting their mental health.

**Statement of Significance:** The present study is the first to evaluate the different effect of pandemic-related remote working on sleep health/habits depending on the chronotype. We found longer sleep duration and improved sleep disturbances among evening-type subjects when working from home. This outcome could be ascribable to a better alignment between the endogenous circadian phase and the working schedule, as remote workers reported later bedtimes and get-up times. Moreover, we showed how improved sleep weakened the susceptibility to depressive symptomatology of evening-type people, highlighting an intriguing implication of working remotely on mental health in this category. Morningness-eveningness predisposition should be considered when designing remote working policies, in order to promote sleep and mental health of late chronotypes during the pandemic emergency, as well as in the post-covid era.

## Introduction

Since the first months of 2020, the COVID-19 pandemic has deeply affected the everyday life of the world population. After a summer period of reduced contagion and death rates, Winter 2020 was marked by a new exacerbation of the pandemic emergency.^1^ The labor market was deeply affected by the emergency period as millions of workers were subjected to exceptional measures worldwide. The most widespread way to cope with the pandemic crisis has been a rapid transition to the remote work modality. According to a recent Eurofound report,^2^ there was an upsurge in teleworking across all European countries during the COVID-19 pandemic. Approximately 40% of the European workforce began to work from home full-time. Similarly, in the United States, 35% of the population shifted from commuting to working remotely.^3^

Notwithstanding the large-scale nature of the remote working implementation, the consequences on sleep health of this unprecedented situation have been scarcely addressed. Remote working removed the need to spend time commuting between home and work and guaranteed greater flexibility of working hours. This situation allowed a better organization of the daily activity, with obvious repercussions on sleep schedule.^4–7^ Consistently, we recently reported a beneficial effect of working from home on sleep quality, insomnia symptoms, and sleep duration among a large sample of the Italian population during the first contagion wave of COVID-19.^6^ A positive effect of the remote working transition on sleep quality and duration was also documented by other investigations.^4,5,8,9^ However, some reports suggested that sleep problems could worsen within the increasing number of people working from home.^10,11^

In our modern society, the issue of the misalignment between the daily social/working schedule and the internal biological clock is a long-standing controversy.^12,13^ In 2006, Wittmann and colleagues^12^ coined the term “*social jetlag*” to give a face to this phenomenon. Consistent evidence pointed to a reduction of the *social jetlag* among the general population in the pandemic period, during which weaker social and working obligations led to a loosening of rigorous sleep/wake schedules.^4,5,9,14,15^ Remarkably, *social jetlag* is intrinsically linked with the circadian typology, being typically more pervasive in the evening chronotype (the so-called “*owls*”). Among this group of people, who tend to go to bed and wake up later in a free-living condition, the mismatch between the endogenous biological and the exogenous social clock is the most pronounced.^13^ This scenario configured an accumulated sleep debt and more sleep problems during the working days in the evening chronotype.^16–18^

An adequate quantity/quality of sleep is crucial for emotional regulation^19,20^ and to preserve mental health,^21,22^ and an extensive literature supported a determining role of both sleep disturbances and short sleep duration in the onset and exacerbation of depressive symptoms.^23–27^ In this vein, it is unsurprising that the evening chronotype has been systematically associated with a mood disturbance propensity.^16,28^ Indeed, several recent reports suggested a causal role of sleep problems in accounting for the association between eveningness and depression.^29–34^ On the other hand, people tending to go to bed and wake up earlier (“*larks*”) are less affected by *social jetlag*, having their sleep-wake rhythms aligned with the common social clock. This situation results in less severe sleep problems and depression symptoms among morning-type people.^16^

The large-scale transition toward remote working during the pandemic represented an unprecedented open-air laboratory to study the relationship between chronobiology and sleep health in a naturalistic environment. The current period emerges as an ideal context to address whether a more flexible working routine could affect sleep quality/quantity of the different circadian typologies and influence the mediating role of sleep between chronotype and depression.

In the present study, we investigated the effect of working from home during the second wave of the COVID-19 outbreak (28 November–11 December 2020) on sleep health/habits of almost nine hundred Italian workers placed along the morningness-eveningness continuum. We evaluated the moderator effect of the working modality (office vs. remote working) on the relationship between chronotype and sleep quality, insomnia symptoms, and sleep duration. We expected to confirm the well-known propensity of the evening-type people to experience sleep problems within the office working group. Meanwhile, we hypothesized that working from home could be associated with specific sleep benefits among the *owls*, reducing the difference of sleep disturbances among the different circadian typologies.

Finally, considering the causal role of sleep disturbances and duration in depressive symptomatology, we investigated the mediation role of sleep in the relationship between chronotype and depression symptoms, evaluating putative differences between office and remote workers. We expected to confirm a significant role of sleep disturbances/duration in accounting for the higher vulnerability to depression of the evening-typology among the office workers. On the other hand, we hypothesized that the mediation effect of sleep could be weakened in the group who worked from home.

## Methods

### Participants and procedure

Data reported in the present study are referred to the group of workers (N=875; mean age ± standard deviation, 36.93 ± 10.57 yrs; range, 20–76 yrs; 729 females) which belonged to an overall sample of 2013 Italian citizens participating in a web-based survey held during the second contagion wave of COVID-19 (28 November– 11 December 2020). The data collection procedure has been documented in detail elsewhere.^35^

The selected sample comprised 610 office workers (35.47 ± 10.17 yrs; 20–68 yrs; 515 females) and 265 full-time remote workers (40.31 ± 10.69 yrs; 23–76 yrs; 214 females). In the present study, we reported demographic data (age, gender), working modality (office working, remote working), and the evaluation of chronotype, sleep quality, insomnia symptoms, and depressive symptomatology, performed through validated questionnaires [Morningness-Eveningness Questionnaire-reduced version (MEQr^36^), Pittsburgh Sleep Quality Index (PSQI^37^), Insomnia Severity Index (ISI^38^), Beck Depression Inventory-second edition (BDI-II^39^).

The Institutional Review Board of the University of L’Aquila approved the study (protocol n. 43066/2020). Online informed consent was obtained from all participants.

### Questionnaires

The MEQr is a 5-item questionnaire to assess the chronotype within the morningness-eveningness continuum. A higher total score (range, 4–25) is interpreted as a tendency to morningness and *vice versa* for eveningness. Cut-off scores are available to classify the chronotype groups (evening-type: 4–10; neither-type: 11–18; morning-type: 19–25). The PSQI is a widely used questionnaire to measure sleep quality. It comprised 19 items, and a higher total score indicates poorer sleep quality (range, 0–21). From the PSQI, we further extracted the answers to the items “sleep duration” (min), “bedtime” (hh:mm), and “get-up time” (hh:mm), which were used in the analyses described in the next paragraph and in the “Supplemental material” section. The ISI is a 7-item instrument to evaluate the severity of insomnia symptoms, where higher scores point to more severe insomnia symptoms (range, 0–28). The BDI-II is a screening instrument to assess the severity of clinical depression. A higher score ranging between 0 and 64 demonstrates more severe depression symptoms.

### Statistical analysis

We performed descriptive statistics on all variables for the two working modality groups (office working, remote working). We evaluated putative differences in gender composition between the two groups through *Chi*-square test. Moreover, we compared office and remote working groups on age and questionnaire scores (MEQr, PSQI, ISI, BDI-II), sleep duration (min), bedtime (hh:mm), and get-up time (hh:mm), using Mann-Whitney *U* test, considering the violation of normality/heteroscedasticity assumptions. All tests were two-tailed, and a *p*-value < 0.05 was considered significant.

According to the research hypotheses, three moderated mediation analyses were run using model 7 of PROCESS macro (version 3.5) for SPSS (version 22.0).^40^ Model 7 assumes that the first stage of the mediation model is moderated. We included MEQr score as independent variable, each sleep outcome [PSQI and ISI score, sleep duration (min)] as individual mediator, and BDI-II score as dependent variable. All the above outcomes were analyzed as continuous variables. Working modality factor (office working, remote working) was assumed as a moderator of the first path of the mediation models (chronotype → sleep variables), and it was entered as a dichotomous dummy variable (office working: 0, remote working: 1). Finally, since prior research indicated that sleep problems and depressive symptoms correlate with age and gender,^41–45^ we included them in the models as continuous and dummy coded (female: 0, male: 1) covariates, respectively. A summary of the three theoretical models tested is provided in Figure 1. Simple slope analyses were performed to explore the nature of the significant interactions between working modality (office working, remote working) and chronotype in predicting sleep variables (sleep quality, insomnia symptoms, sleep duration). The statistical significance of the conditional indirect effects was ascertained by means of 5,000 bootstrap samples to create bias-corrected confidence intervals (*CIs*: 95%) with heteroscedasticity-consistent standard errors (SEs). Moderated mediation models were considered significant and accepted when the interval between the 95% bootstrapped lower limit (*BootLLCI*) and upper limit of CIs (*BootULCI*) of the index of moderated mediation (the difference between conditional indirect effects) does not contain 0. Finally, to further clarify how remote working affected the sleep schedule within the morningness-eveningness continuum, we analyzed how chronotype scores interact with working modality in predicting sleep onset and offset time. Therefore, two explorative moderation models were tested, assuming “working modality” as moderator of the effect of chronotype on bedtime and get-up time (see “Supplemental materials”).

**Figure 1.**
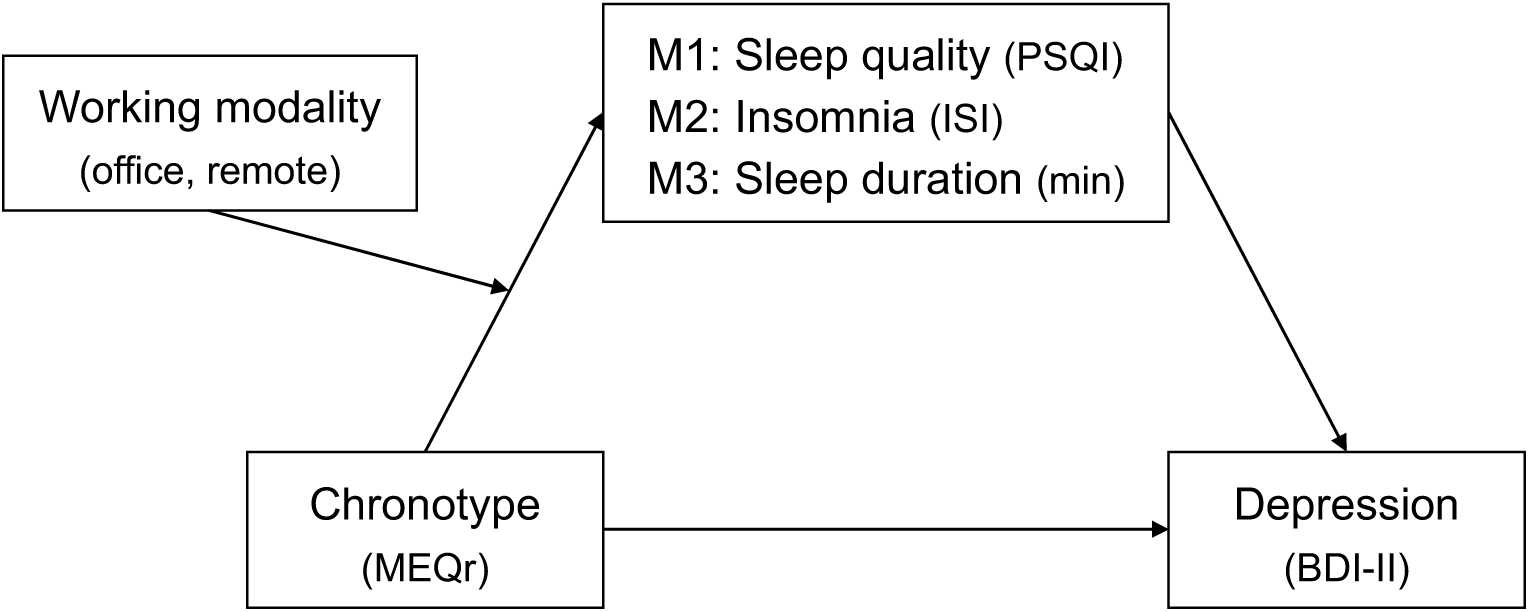
The three theoretical moderated mediation models tested (M1, M2, M3). Three mediators (sleep quality, insomnia symptoms, sleep duration) are hypothesized to mediate the relationship between morningness-eveningness continuum and severity of depression symptoms in a context where working modality (office working, remote working) moderate the effect of chronotype on sleep variables. Each model was adjusted for age and gender. *Abbreviations:* PSQI, Pittsburgh Sleep Quality Index; ISI, Insomnia Severity Index; MEQr, Morningness-Eveningness Questionnaire-reduced version; BDI-II, Beck Depression Inventory-second edition.

## Results

### Characteristics of participants

Demographic composition of the two groups (office work, remote work) and descriptive statistics of the main study variables are shown in Table 1.

**Table 1.** Characteristics of participants divided by working modality (office, remote). Results of the comparisons between the working modality groups are also shown.

|  | Working modality |  | Statistic | df | p |
| --- | --- | --- | --- | --- | --- |
|  | Office<br>[N = 610 (69.7%)] | Remote<br>[N = 265 (30.3%)] |  |  |  |
|  | Mean ± SD | Mean ± SD |  |  |  |
| Gender |  |  |  |  |  |
| Male | 95 (15.6%) | 51 (19.2%) | 1.791* | 1 | 0.181 |
| Female | 515 (84.4%) | 214 (80.8%) |  |  |  |
| Age | 35.467 ± 10.174 | 40.309 ± 10.694 | 58326<sup>†</sup> | 873 | <.001 |
| MEQr score | 15.867 ± 3.494 | 15.430 ± 3.850 | 76014.5<sup>†</sup> | 873 | 0.160 |
| PSQI score | 6.693 ± 3.504 | 7.015 ± 3.598 | 76367.5<sup>†</sup> | 873 | 0.192 |
| ISI score | 7.428 ± 5.268 | 7.815 ± 5.356 | 77398<sup>†</sup> | 873 | 0.318 |
| Sleep duration (min) | 403.365 ± 66.462 | 401.624 ± 64.434 | 79169<sup>†</sup> | 873 | 0.626 |
| Bedtime (hh:mm) | 23:11 ± 1:05 | 23:37 ± 1:16 | 63901<sup>†</sup> | 873 | <.001 |
| Get-up time (hh:mm) | 7:13 ± 1:03 | 7:41 ± 1:09 | 56625<sup>†</sup> | 873 | <.001 |
| BDI-II score | 11.141 ± 8.620 | 11.660 ± 9.698 | 79897<sup>†</sup> | 873 | 0.787 |

\* Chi-square, <sup>†</sup> Mann-Whitney U.
Abbreviations: SD, standard deviation; df, degrees of freedom; PSQI, Pittsburgh Sleep Quality Index; ISI, Insomnia Severity Index; MEQr, Morningness-Eveningness Questionnaire-reduced version; BDI-II, Beck Depression Inventory-second edition.

The two samples did not differ in MEQr, PSQI, ISI, and BDI-II scores, as well as in sleep duration and gender proportion. However, the office working group was significantly younger and reported earlier bedtime and get-up time.

### Moderated mediation analyses

The regressions on mediators (PSQI score, ISI score, sleep duration) were significant for each model (Model 1: *R*^*2*^ = .069, *F* = 12.856, *p* < .001; Model 2: *R*^*2*^ = .063, *F* = 11.617, *p* < .001; Model 3: *R*^*2*^ = .094, *F* = 18.057, *p* < .001). Likewise, the regressions on BDI-II scores were significant for all the models (Model 1: *R*^*2*^ = 0.289, *F* = 88.123, *p* < .001; Model 2: *R*^*2*^ = 0.373, *F* = 129.62, *p* < .001; Model 3: *R*^*2*^ = 0.135, *F* = 33.947, *p* < .001). As showed in Table 2, older age was associated with lower sleep quality, more severe insomnia, shorter sleep duration, and lower depressive symptoms in all the models. Male subjects reported better sleep quality and less severe insomnia symptoms than females, while no difference in sleep duration between genders emerged. Men showed a lower severity of depression in each model.

**Table 2.** Unstandardized effects (*B*), *t*-value, and significance of the covariates (age, gender) for the three models, including sleep quality (PSQI score; Model 1), insomnia symptoms (ISI score; Model 2), and sleep duration (min; Model 3) as mediators.

| **Covariate effects** | ***B*** | ***t*** | ***p*** |
| --- | --- | --- | --- |
| **Model 1** |  |  |  |
| Age → PSQI | .051 | 4.480 | <.001 |
| Gender* → PSQI | -1.230 | -3.915 | <.001 |
| Age → BDI-II | -2.188 | -3.120 | .002 |
| Gender* → BDI-II | -.067 | -2.690 | .007 |
| **Model 2** |  |  |  |
| Age → ISI | .052 | 3.040 | .002 |
| Gender* → ISI | -1.943 | -4.112 | <.001 |
| Age → BDI-II | -.054 | -2.336 | .019 |
| Gender* → BDI-II | -1.820 | -2.764 | .006 |
| **Model 3** |  |  |  |
| Age → Sleep duration | -1.834 | -8.752 | <.001 |
| Gender* → Sleep duration | -6.908 | -1.196 | .232 |
| Age → BDI-II | -.071 | -2.493 | .013 |
| Gender* → BDI-II | -3.998 | -5.217 | <.001 |

\* Female was used as reference for "Gender" factor.
*Abbreviations:* PSQI, Pittsburgh Sleep Quality Index; ISI, Insomnia Severity Index; BDI-II, Beck Depression Inventory-second edition.

As reported in Table 3, both the MEQr scores and the sleep variables (PSQI score, ISI score, sleep duration) were significantly associated with the BDI-II scores in all the models (direct effects). Tendency to eveningness, lower sleep quality, more severe insomnia, and shorter sleep duration predicted greater depression symptoms. The conditional direct effects at the value of the moderator (office working, remote working) indicated that the tendency to morningness was associated with better sleep quality, lower severity of insomnia symptoms, and longer sleep duration in the office working group. On the other hand, no significant relationship between chronotype and sleep variables emerged among the remote workers.

**Table 3.** Direct effects and conditional direct effects at the value of the moderator (office working, remote working) for the three models, including sleep quality (PSQI score; Model 1), insomnia symptoms (ISI score; Model 2), and sleep duration (min; Model 3) as mediators.

| Direct effects | *B* | *t* | *p* |
| --- | --- | --- | --- |
| **Model 1** |  |  |  |
| MEQr → BDI-II | -.274 | -3.724 | <.001 |
| PSQI → BDI-II | 1.245 | 16.613 | <.001 |
| **Model 2** |  |  |  |
| MEQr → BDI-II | -.233 | -3.376 | <.001 |
| ISI → BDI-II | .971 | 20.775 | <.001 |
| **Model 3** |  |  |  |
| MEQr → BDI-II | -.445 | -5.560 | <.001 |
| Sleep duration → BDI-II | -.038 | -8.535 | <.001 |
| **Conditional direct effects** |  |  |  |
| **Model 1** |  |  |  |
| Office working: MEQr → PSQI | -.247 | -6.151 | <.001 |
| Remote working: MEQr → PSQI | -.101 | -1.825 | .068 |
| **Model 2** |  |  |  |
| Office working: MEQr → ISI | -.374 | -6.200 | <.001 |
| Remote working: MEQr → ISI | -.141 | -1.696 | 0.090 |
| **Model 3** |  |  |  |
| Office working: MEQr → Sleep duration | 3.023 | 4.102 | <.001 |
| Remote working: MEQr → Sleep duration | 0.151 | .149 | 0.881 |

*Abbreviations:* PSQI, Pittsburgh Sleep Quality Index; ISI, Insomnia Severity Index; MEQr, Morningness-Eveningness Questionnaire-reduced version; BDI-II, Beck Depression Inventory-second edition.

The “working modality” moderator was significant in each model (Table 4), indicating that remote workers reported higher sleep quality, lower insomnia symptoms, and longer sleep duration than the office working group. The interaction between MEQr scores and the working modality moderator was significant in each model, suggesting a different linear relationship between chronotype and sleep variables comparing the office and remote working groups (Figure 2).

**Table 4.** Moderator and interaction effects for the three models, including sleep quality (PSQI score; Model 1), insomnia symptoms (ISI score; Model 2), and sleep duration (min; Model 3) as mediators.

| **Moderator effects** | ***B*** | ***t*** | ***p*** |
| --- | --- | --- | --- |
| **Model 1** |  |  |  |
| Working modality* → PSQI | −2.242 | −2.062 | **.039** |
| **Model 2** |  |  |  |
| Working modality* → ISI | −3.555 | −2.175 | **.030** |
| **Model 3** |  |  |  |
| Working modality* → Sleep duration | 53.026 | 2.654 | **.008** |
| **Interaction effects** |  |  |  |
| **Model 1** |  |  |  |
| Working modality* X MEQr → PSQI | .146 | 2.158 | **.031** |
| **Model 2** |  |  |  |
| Working modality* X MEQr → ISI | .233 | 2.291 | **.021** |
| **Model 3** |  |  |  |
| Working modality* X MEQr → Sleep duration | −2.872 | −2.309 | **.012** |

\* Office working was used as reference for “Working modality” factor.
*Abbreviations:* PSQI, Pittsburgh Sleep Quality Index; ISI, Insomnia Severity Index; MEQr, Morningness-Eveningness Questionnaire-reduced version.

**Figure 2.**
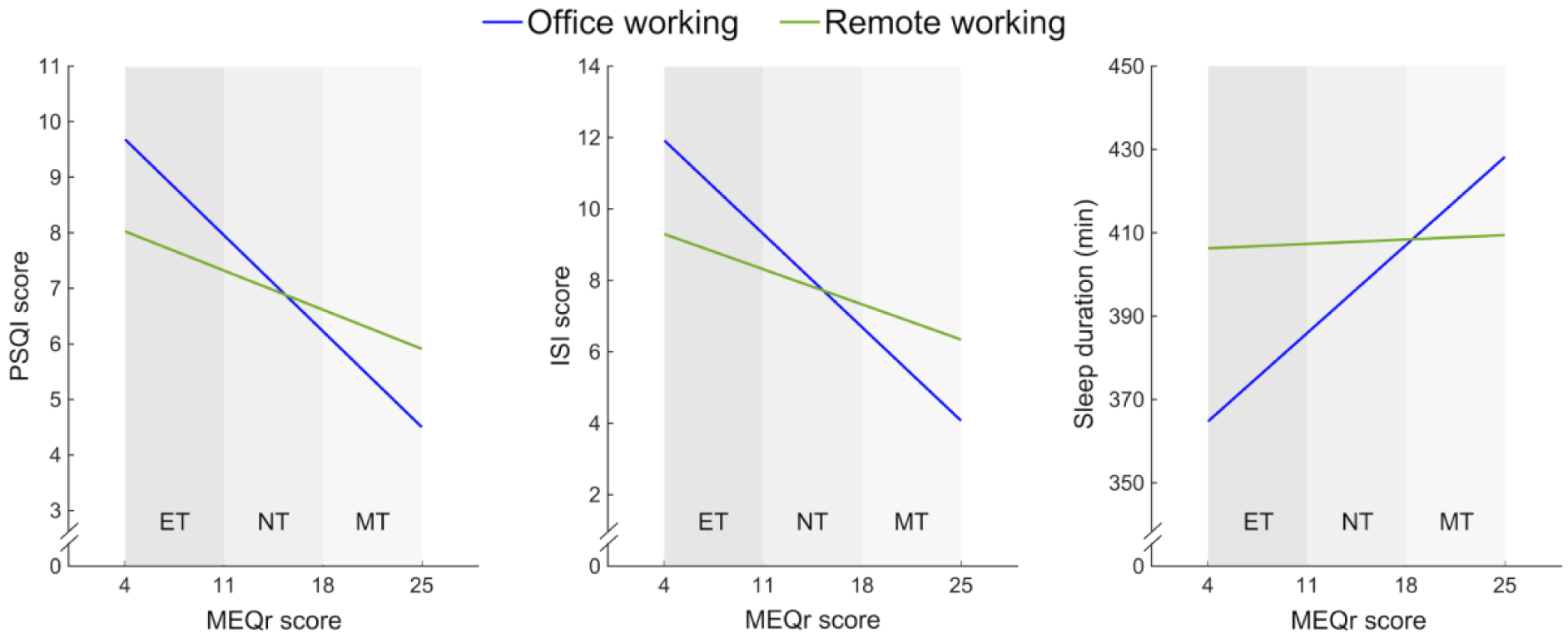
Simple slope analyses of the interaction between MEQr scores and working modality [office working (blue line), remote working (green line)] on sleep quality (PSQI score), insomnia symptoms (ISI score), and sleep duration (min). Gray bands discriminate chronotypes according to the validated cut-off scores. *Abbreviations:* ET, evening type; NT, neither type; MT, morning type; PSQI, Pittsburgh Sleep Quality Index; ISI, Insomnia Severity Index; MEQr, Morningness-Eveningness Questionnaire-reduced version.

Finally, as reported in Figure 3, all the conditional indirect effects were significant for the office working group, indicating that the sleep variables partially mediated the effect of chronotype on depression symptoms. On the other hand, no significant indirect effect was detected in the remote working group. Consistently, the index of moderated mediation was significant in each model (Model 1: 0.182 [0.008, 0.361]; Model 2: 0.227 [0.031, 0.430]; Model 3: 0.110 [0.015, 0.221]), indicating that working from home suppressed the mediation effect of sleep variables on the association between chronotype and depression.

**Figure 3.**
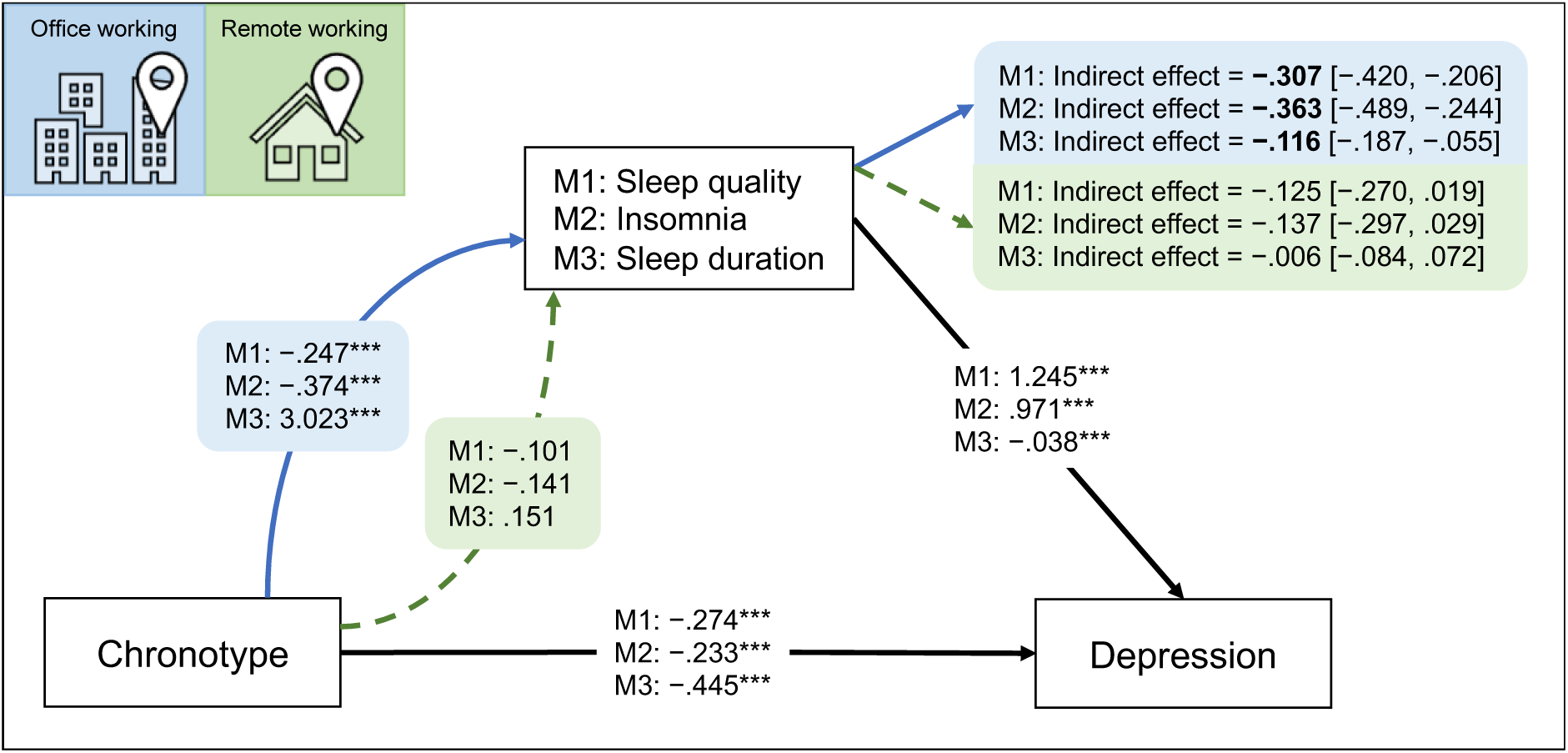
Summary of the results of the three moderated mediation models (M1, M2, M3). The figure reports the unstandardized coefficients of direct effects, conditional direct effects at the value of moderator, and conditional indirect effects with bootstrapped computed confidence intervals for the two levels of moderator [office working (blue arrow/area), remote working (green arrow/area)]. Significance is indicated with asterisks (\*\*\**p* < .001) and bold characters for direct and indirect effects, respectively.

## Discussion

The COVID-19 emergency pervasively impacted the daily routine of millions of workers worldwide. Consistent with European and American reports during the pandemic,^2,3^ three out of ten individuals of our sample worked full-time from home. In line with recent investigations,^4–7^ the remote working group showed a general delay of bedtime and get-up time. We hypothesized that evening-type subjects could have benefited from such scenario, as their sleep time was better aligned with the endogenous circadian phase than in the office working condition. The analyses confirmed our prediction: the tested models revealed a significant interaction between chronotype scores and working modality (office, remote) in predicting sleep variables. Remarkably, we demonstrated that the well-documented relationship between chronotype and sleep problems/duration^16–18^ was limited to the office working group. Therefore, our findings suggest that the propensity to sleep problems and shorter sleep duration of evening-type people did not depend on chronotype *per se*, and the eveningness represented a risk factor only in the office working condition.

The outcomes of the additional models (see “Supplemental materials”) contributed to further clarifying the pattern of results. We showed that working from home impacted the relationship between chronotype and get-up time but did not affect the association with bedtime. In the remote working group, eveningness was related to a stronger tendency to get-up later than the office working sample. On the other hand, the inclination of evening-type people to go to bed later than morning-type was similar in the two working modality groups. Therefore, later bedtimes were not adequately compensated by later get-up times when participants had to reach the workplace, giving rise to an accumulated sleep debt among the late chronotypes, as confirmed by their tendency to shorter sleep duration. On the other hand, the strengthening of the association between get-up times and chronotype scores implies that the *owls* slept more when they worked from home, leading to the extinction of the sleep duration differences between circadian typologies.

Our findings are consistent with studies showing that chronotype modulates the effect of the working schedule on sleep patterns.^46,47^ Late chronotypes are marked by shorter sleep duration and more severe sleep disturbances compared with early ones when working in the morning.^46^ Consistently, a chronotype-based working routine was associated with increased sleep duration and quality by reducing the *social jetlag* among the evening-type population.^47^

Several studies demonstrated a positive effect of working from home on sleep patterns during the current pandemic.^4,5,7–9^ This literature is consistent with investigations on the student population under remote learning due to the COVID-19 emergency, where participants delayed the sleep time and reported longer sleep duration and improved sleep quality.^48–51^ Notwithstanding the lack of an evaluation of possible differential effects as a function of circadian typology in these studies, the results were consistently interpreted as a general tendency to synchronize the sleep/wake cycle with the individual biological clock when daily schedules are less strongly dictated by the office/school hours. Interestingly, we failed to highlight any difference in mean sleep quality, sleep duration, and severity of insomnia symptoms in the preliminary direct between-group comparisons. However, the beneficial effect of remote working on sleep emerged by including the interaction between chronotype scores and working modality in the models. This evidence could account for some of the inconsistencies in the literature addressing the effect of remote working during the pandemic period.^10,11^ The individual circadian preference could act as a confounding variable, resulting in misleading conclusions when studying the consequences of the working modality on sleep health. Therefore, we caution that future studies in this field duly consider chronotype and its interaction with both working modality and schedule.

As far as the depressive symptomatology is concerned, we confirmed the tendency of the evening-type participants to experience more severe symptoms,^16,28^ as well as the well-documented positive relationship between both sleep problems and short sleep duration and more severe depression symptoms.^23–27^ Meanwhile, the loosening of the association between sleep disturbances/duration and chronotype in the group who worked from home corroborated the second goal of this study: determining whether remote working affected the mediation role of sleep between circadian typologies and depression symptoms. The three moderated mediation models demonstrated that poorer sleep quality, more severe insomnia symptoms, and shorter sleep duration partially explained the tendency of evening chronotype to experience depression only among people who had to reach the workplace. This outcome is consistent with the growing literature supporting a causative role of sleep disturbances and shorter sleep duration in explaining the eveningness susceptibility to depressive symptomatology.^29–34^ On the other hand, we demonstrated that the sleep-dependent vulnerability to depression of late chronotypes disappeared under remote working. Therefore, the improvement of sleep problems while working from home resulted in an indirect benefit on the mental health of evening-type participants, relieving their predisposition to depressive symptoms.

The results of this study solicit a discussion at the community level. Our modern society forces all employees to fit a “standard” work schedule typically oriented to morningness. Social pressure imposes to get up early in the morning since the school period,^52^ notwithstanding that the early morning activities are associated with sleep problems and fatigue among the general population.^53^ This situation is even more pronounced when the large and intrinsic variability in the biological circadian predispositions is considered,^46,47^ configuring a latent penalization of evening-type people. Considering the individual circadian predisposition in managing the working environment could promote the sleep and mental health of evening-type individuals. The vaccination campaign and the gradual mitigation of the pandemic crisis are leading workers to resume their pre-pandemic daily routine worldwide. In this vein, our results have large-scale implications spanning to the post-pandemic period, considering that circadian predisposition has a substantial genetic component^54,55^ which as such could be hardly manipulated, and the current literature estimated a 10–20% prevalence of *owls* among the adult population,^6,56–60^ which is also higher among the young people^16,61^. The outcomes of the present study should be taken into account when designing remote working policies during the current pandemic, as well as in the post-covid era.

## Limitations

Our pattern of results was obtained in a large sample of workers. Moreover, the inclusion of demographic factors (age and gender) in all the tested models confuted the possibility that the younger age of the office working group could have biased our results. However, some limitations should be acknowledged. We adopted a cross-sectional design, and the sample comprised a higher prevalence of women. Moreover, our findings relied on regression analyses, so that the direction of the effects could be only hypothesized. A longitudinal analysis might confirm our results clarifying the causal relationship between the investigated variables. Additionally, chronotype and sleep variables were assessed using self-reported questionnaires, and the evaluation of sleep habits did not discriminate between workdays and free days. However, a sleep assessment targeted on workdays could have provided even stronger evidence of the interaction between chronotype and working modality. Moreover, it is to be noted that PSQI scores predominantly reflect sleep quality/patterns of workdays.^62^ Finally, an *ad hoc* evaluation of the *social jetlag* phenomenon through, e.g., the Munich Chronotype Questionnaire,^63^ might have contributed to better understanding the effect of working from home during this unprecedented pandemic period.

## Data Availability

The data underlying this article will be shared on reasonable request to the corresponding author.

## Acknowledgments

Conceptualization, F.S. and M.F.; Methodology, F.S.; Investigation, F.S., G.A., L.V.; Data curation, F.S.; Formal analysis, F.S.; Writing – original draft, F.S.; Writing – review & editing, F.S., A.D.A., M.G., S.S., V.A., and M.F.; Supervision, M.F.

## Data Availability Statement

The data underlying this article will be shared on reasonable request to the corresponding author.

