## Supplemental materials for "Working from home due to the COVID-19 pandemic abolished the sleep disturbance vulnerability of late chronotypes relieving their predisposition to depression"

Two additional moderation models were tested using model 1 of PROCESS macro (version 3.5) for SPSS (version 22.0).[40] We assumed that working modality (office working, remote working) moderated the effect of chronotype on bedtime and get-up time (Figure S1). Both the models included the covariance of age and gender.

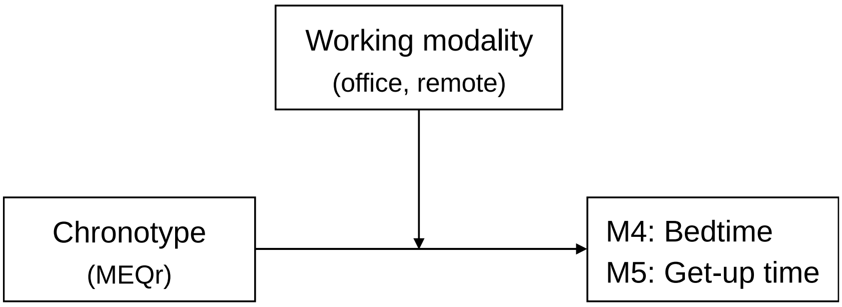


**Figure S1.** The two additional moderation models tested (M4, M5). Chronotype is hypothesized to predict bedtime and get-up time, while the working modality (office working, remote working) moderated this relationship.
*Abbreviations:* MEQr, Morningness-Eveningness Questionnaire-reduced version.

The regressions on bedtime and get-up time were significant (Model 4: $R^2$ = .247, $F$ = 56.852, $p$ < .001; Model 5: $R^2$ = .274, $F$ = 65.685, $p$ < .001). The covariate effect of "gender" emerged significant for the bedtime variable ($B$ = 14.51 min, $t$ = 2.630, $p$ = .009), indicating that female respondents went to bed earlier. Age was a significant predictor of get-up time ($B$ = −1.58 min, $t$ = −8.509, $p$ < .001), indicating that older age was associated with earlier get-up time.

The conditional direct effects at both values of the moderator (office working, remote working) were significant in both the models. In particular, MEQr score predicted significantly bedtime (office working: $B$ = −9.80 min, $t$ = −10.029, $p$ < .001; remote working: $B$ = −7.92 min, $t$ = −11.154, $p$ < .001) and get-up time (office working: $B$ = −8.50 min, $t$ = −9.317, $p$ < .001; remote working: $B$ = −6.05 min, $t$ = −9.221, $p$ < .001), confirming the tendency to delayed sleep time of evening-type people in both the working modality conditions. The "working modality" moderator was significant in both the models (M4: $B$ = 51.55 min, $t$ = 2.682, $p$ = .008; M5: $B$ = 69.27 min, $t$ = 3.874, $p$ = .001), showing that people working from home went to bed and got-up later. Finally, the significant interaction between "working modality" and MEQr score was limited to the get-up time variable (M4: $B$ = −1.87 min, $t$ = −1.564, $p$ = .118; M5: $B$ = −2.30 min, $t$ = −2.130, $p$ = .033), highlighting that the relationship between chronotype and rising time was stronger in those who worked from home than in the office working condition (Figure S2).

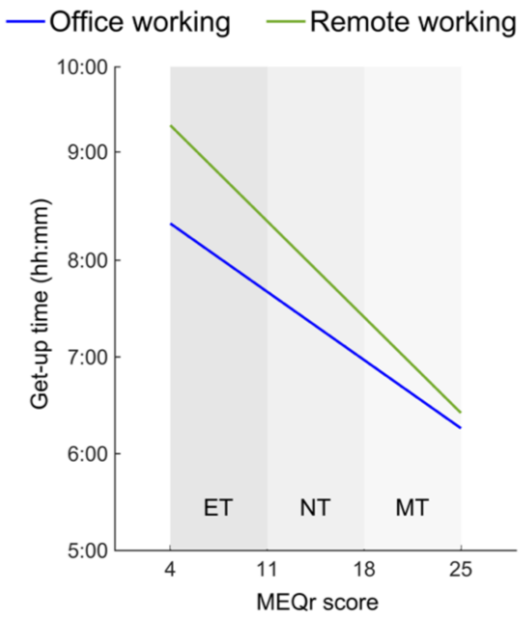


**Figure S2.** Simple slope analysis of the interaction between MEQr scores and working modality [office working (blue line), remote working (green line)] on get-up time (hh:mm). Gray bands identify chronotypes according to the validated cut-off scores.

*Abbreviations:* ET, evening type; NT, neither type; MT, morning type; MEQr, Morningness-eveningness questionnaire-reduced version.
